# Current Understanding of COVID-19 Clinical Course and Investigational Treatments

**DOI:** 10.1101/2020.04.19.20071548

**Authors:** Richard B. Aguilar, Patrick Hardigan, Bindu Mayi, Darby Sider, Jared Piotrkowski, Jinesh P. Mehta, Jenankan Dev, Yelenis Seijo, Antonio Lewis Camargo, Luis Andux, Kathleen Hagen, Marlow B. Hernandez

## Abstract

**Importance:** Currently, there is no unified framework linking disease progression to established viral levels, clinical tests, inflammatory markers, and investigational treatment options.

**Objective:** It may take many weeks or months to establish a standard treatment approach. Given the growing morbidity and mortality with respect to COVID-19, we present a treatment approach based on a thorough review of scholarly articles and clinical reports. Our focus is on staged progression, clinical algorithms, and individualized treatment.

**Evidence Review:** We followed the protocol for a quality review article proposed by Heyn et. al.^1^ A literature search was conducted to find all relevant studies related to COVID-19. The search was conducted between April 1, 2020 and April 13, 2020 using the following electronic databases: PubMed (1809 to present), Google Scholar (1900 to present), MEDLINE (1946 to present), CINAHL (1937 to present), and Embase (1980 to present). Keywords used included *COVID-19, 2019-nCov, SARS-CoV-2, SARS-CoV*, and *MERS-CoV*, with terms such as *efficacy, seroconversion, microbiology, pathophysiology, viral levels, inflammation, survivability*, and *treatment and pharmacology*. No language restriction was placed on the search. Reference lists were manually scanned for additional studies.

**Findings:** Of the articles found in the literature search, 70 were selected for inclusion in this study (67 cited in the body of the manuscript and 3 additional unique references in the Figures).

The articles represent work from China, Japan, Taiwan, Vietnam, Rwanda, Israel, France, the United Kingdom, the Netherlands, Canada, and the United States. Most of the articles were cohort or case studies, but we also drew upon information found in guidelines from hospitals and clinics instructing their staff on procedures to follow. In addition, we based some decisions on data collected by agencies such as the CDC, FDA, IHME, ISDA, and Worldometer. None of the case studies or cohort studies used a large number of participants. The largest group of participants numbered less than 500 and some case studies had fewer than 30 patients. However, the review of the literature revealed the need for individualized treatment protocols due to the variability of patient clinical presentation and survivability. A number of factors appear to influence mortality: the stage at which the patient first presented for care, pre-existing health conditions, age, and the viral load the patient carried.

**Conclusion and Relevance:** COVID-19 can be divided into three distinct Stages, beginning at the time of infection (Stage I), sometimes progressing to pulmonary involvement (Stage II, with or without hypoxemia) and less frequently to systemic inflammation (Stage III). In addition to modeling the stages of disease progression, we have also created a treatment algorithm which considers age, comorbidities, clinical presentation, and disease progression to suggest drug classes or treatment modalities. This paper presents the first evidence-based recommendations for individualized treatment for COVID-19.

**Key Points:** *Question:* What are the most effective treatment recommendations for COVID-19?

*Findings:* COVID-19 can be divided into three distinct Stages, beginning at the time of infection (Stage I), sometimes progressing to pulmonary involvement (Stage II, with or without hypoxemia) and less frequently to systemic inflammation (Stage III). In addition to modeling the stages of disease progression, we also created a treatment algorithm which considers age, comorbidities, clinical presentation, and disease progression to suggest drug classes or treatment modalities.

*Meaning:* This paper presents the first evidence-based recommendations for individualized treatment for COVID-19.

## Introduction

The coronavirus disease 2019 (COVID*-*19) pandemic is raging throughout the globe. In the United States alone, as of April 10, 2020, there were 560,933 cases along with 22,127 deaths (Centers for Disease Control and Prevention, CDC).^2^ A mathematical model created by The Institute for Health Metrics and Evaluation (IHME) predicts that in the United States, assuming continued full social distancing, the number of new cases will peak in late April, and related deaths will peak by mid-to-late May.^3^ This creates a critical and immediate need for medical treatment and resources.

Preliminary data in the US suggests that COVID*-*19 may be more infectious and lethal than Influenza H1N1. In the general population, current case-fatality rates for COVID*-*19 are about 3.9%, and infection rates are about 2.5 under normal conditions.^4^ To place this in context, Figure 1 provides a comparison of the reproduction rate and case-fatality rates for major respiratory virus pandemics.^5-7^ Data strongly emphasizes early intervention to reduce case-fatality and inhibit reproductive rates.

**Figure 1.**
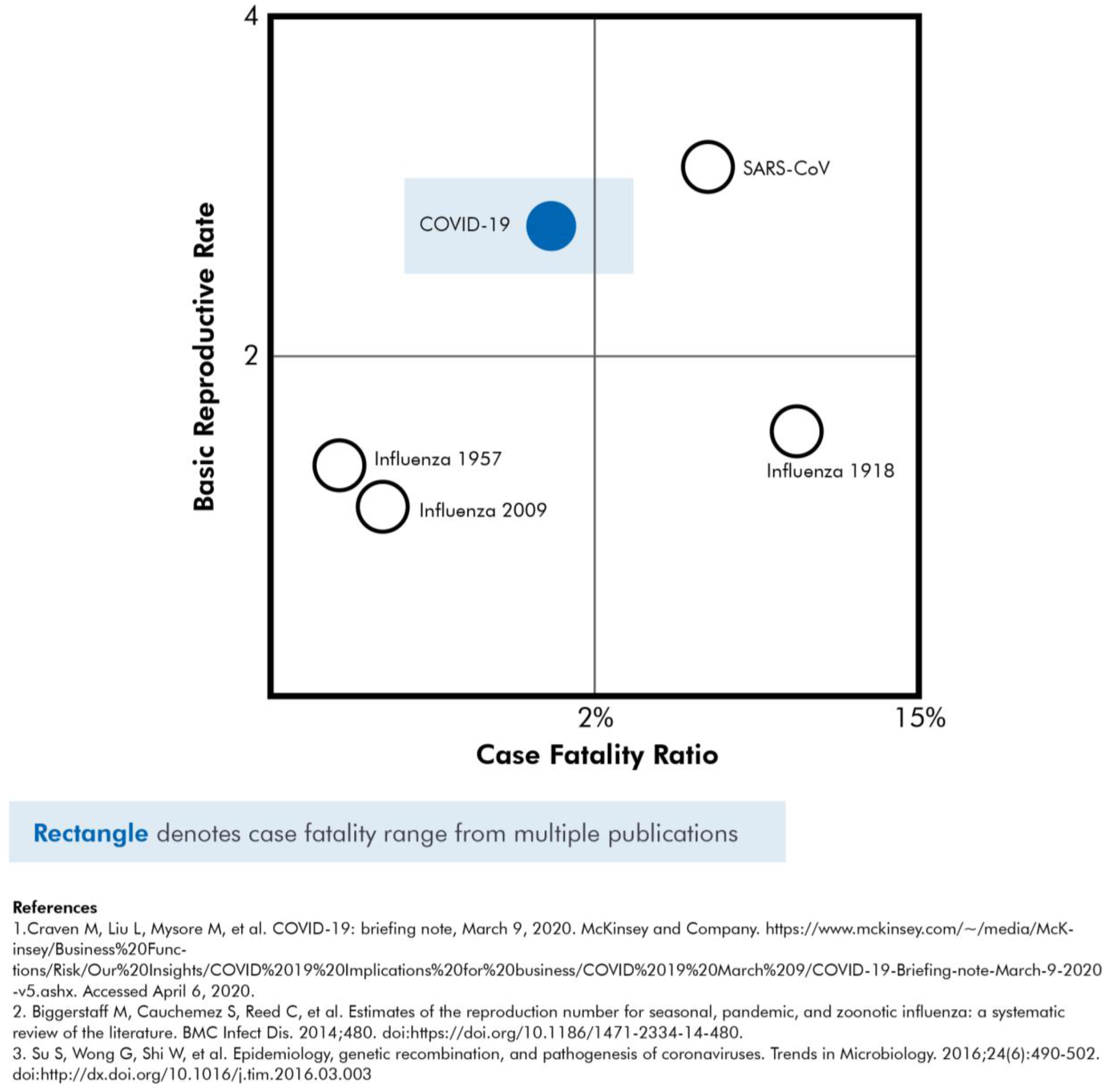
Reproduction Rate and Case-Fatality Rates for Major Respiratory Virus Pandemics

A number of articles have been published on the clinical course and treatment of the disease.^8-10^ The majority of patients present with more than one symptom on admission, although the combination of fever, cough, and shortness of breath is rare. Siddiqi and Mehra proposed a staged progression model based on observed clinical courses in published studies.^11^ In Stage 1, or the mild phase, the virus multiplies and establishes residence in the host, predominantly in the respiratory tract. In Stage 2, we see that viral multiplication and localized inflammation in the lungs is common. Stage 3 is marked by extra-pulmonary systemic hyperinflammation syndrome. The prognosis and recovery from Stage 3 is generally poor. Rapid recognition of what stage the patient is in and deployment of appropriate therapy may have the greatest yield.

Common correlates with poorer outcomes include age, hypertension, diabetes, coronary artery disease, chronic lung disease, and malignancies.^12^ Research also finds variations in outcomes due to a dysregulated and exuberant immune response. Patients requiring intensive care have significantly higher levels of IL-6, CRP, ferritin, and D-Dimer. An important therapeutic modality may be to downregulate the cytokine storm, particularly in severe illness.^13^ The literature also suggest that disease progression can be predicted. During the severe acute respiratory syndrome (SARS) pandemic, a retrospective analysis revealed that 2-week cumulative case data could help estimate the total case numbers with accuracy – well before the date of the last reported case.^14^

As we have found, there is no unified framework linking disease progression to established viral levels, clinical tests, inflammatory markers, and investigational treatment options. Given that it may take many weeks or months to establish a standard treatment approach and there is a growing morbidity and mortality, we present an initial treatment approach based on a thorough review of currently available scholarly articles and clinical reports. Our focus is on staged progression, clinical algorithms, and individualized treatment.

## Methods

We followed the protocol for a quality review article proposed by Heyn et al.^1^ A literature search was conducted to find all relevant studies related to COVID-19. The search was conducted between April 1, 2020 and April 13, 2020 using the following electronic databases: PubMed (1809 to present), Google Scholar (1900 to present), MEDLINE (1946 to present), CINAHL (1937 to present), and Embase (1980 to present). Keywords used included *COVID-19, 2019-nCov, SARS-CoV-2, SARS-CoV*, and *MERS-CoV*, with terms such as *efficacy, seroconversion, microbiology, pathophysiology, viral levels, inflammation, survivability*, and *treatment and pharmacology*. No language restriction was placed on the search. Reference lists were manually scanned for additional studies. From this systematic review a model was created that incorporated clinical course, diagnostics, disease management, and treatment.

Our results focus on recommendations for individualized treatment, by selecting the most appropriate drug or modality for the patient, carefully weighing risks and benefits. Clinicians and patients should understand the staged progression of COVID-19 (Figure 2). As such, we present a treatment algorithm that recommends no treatment for some and specific treatment for others, depending on age, comorbidities, and symptom severity (Figure 3).

**Figure 2.**
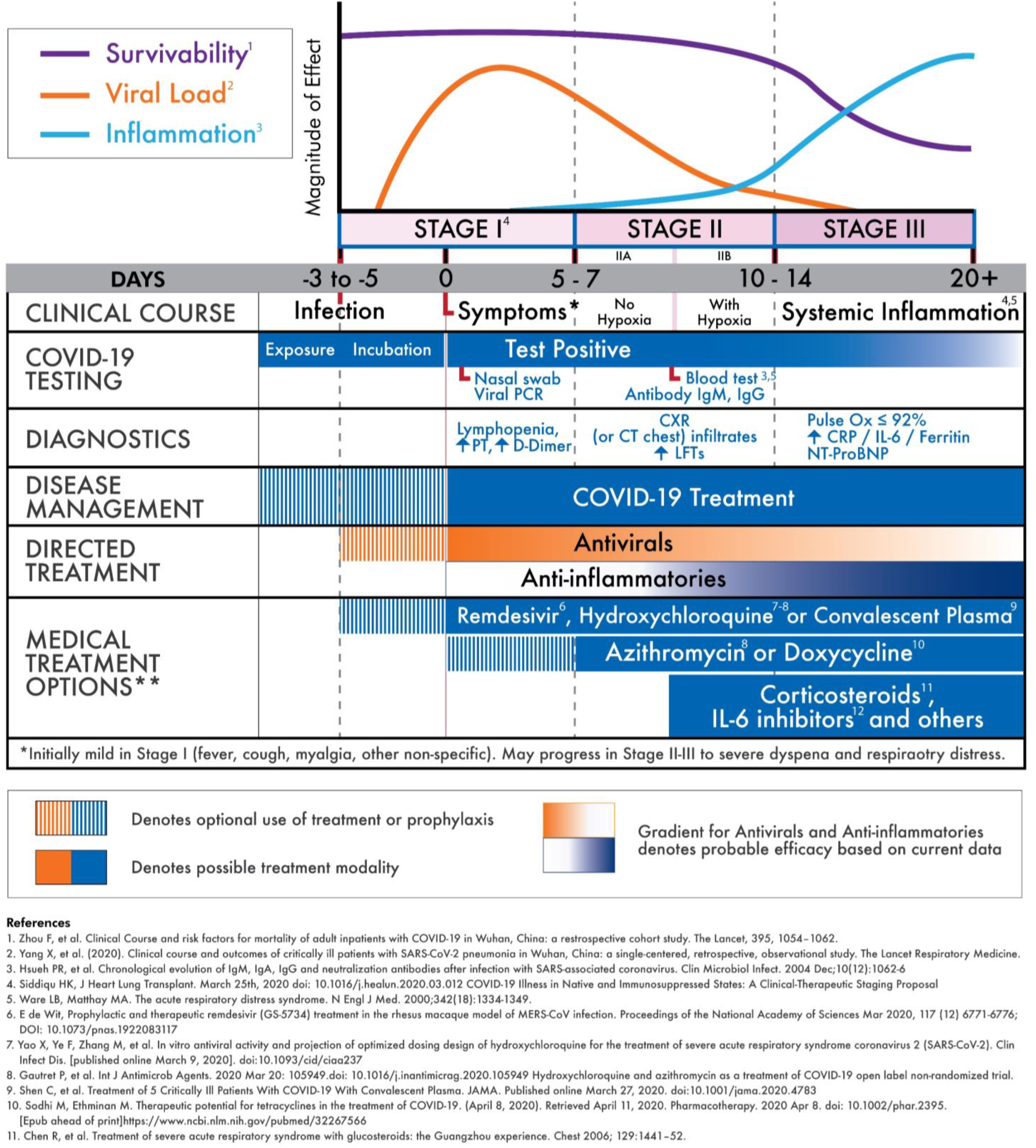
COVID-19 Clinical Stages and Management Strategy

**Figure 3.**
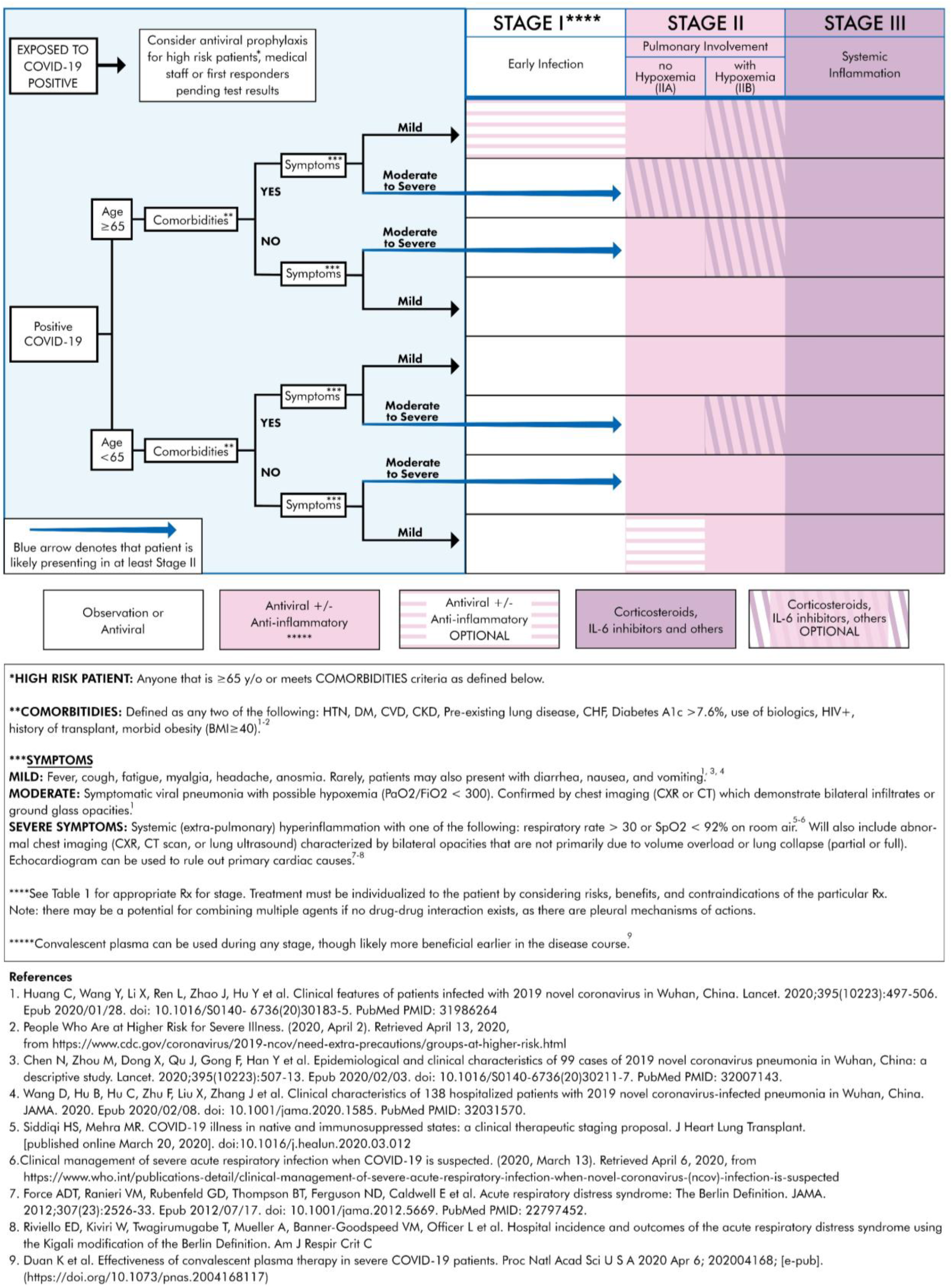
Treatment Algorithm for COVID-19+ Patients Based on Clinical Presentation and Therapeutic Staging

## Results

Based on our thorough review of the literature, we correlated the disease course to COVID-19 testing, diagnostic options, and treatment strategies (see Figure 2). COVID-19 can be divided into three distinct Stages, beginning at the time of infection (Stage I), sometimes progressing to pulmonary involvement (Stage II, with or without hypoxemia) and less frequently to systemic inflammation (Stage III). We also created a treatment algorithm which considers age, comorbidities, clinical presentation, and disease progression to suggest drug classes or treatment modalities (see Figure 3). The specific treatments are summarized in Table 1.^20-64^

**Table 1.**
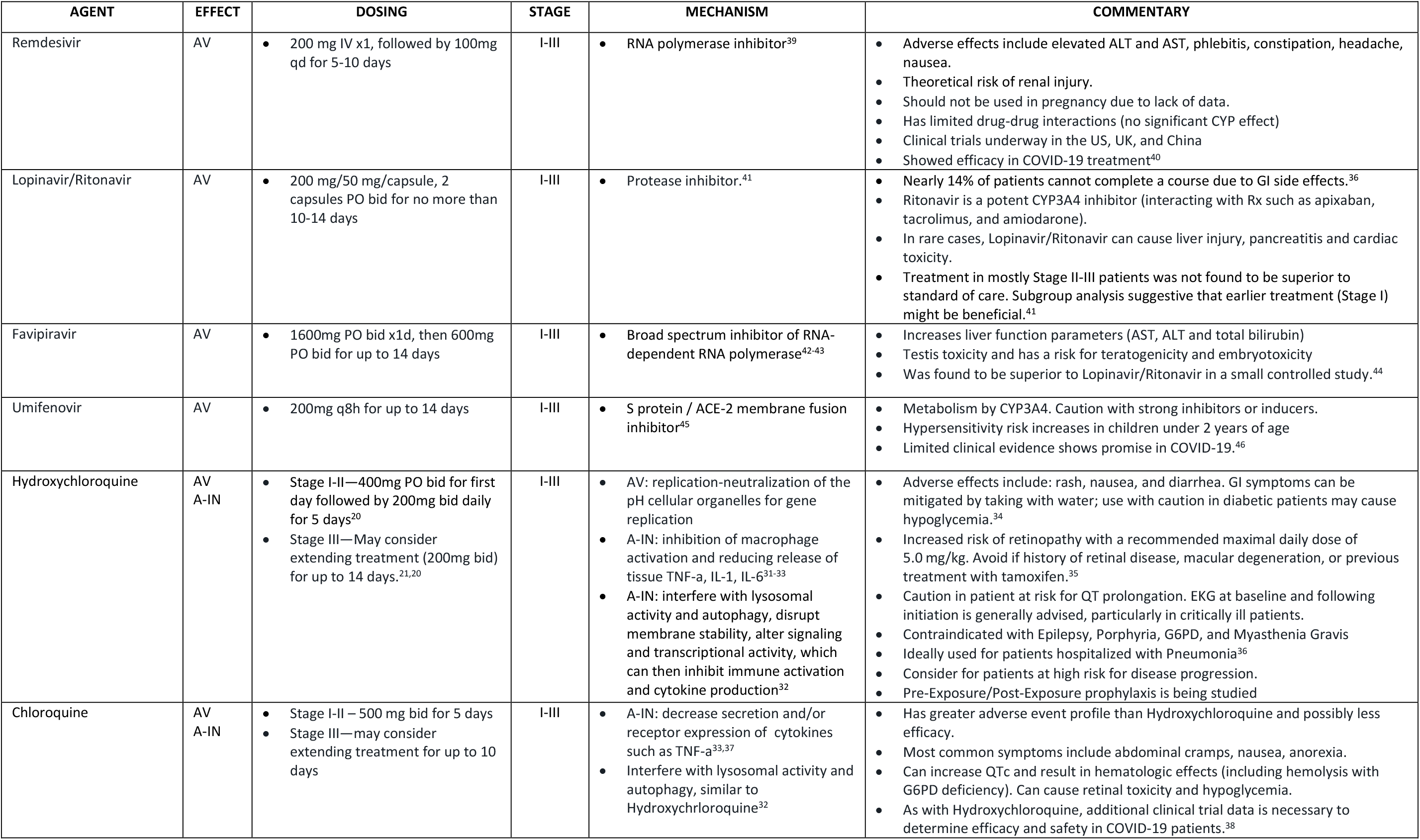
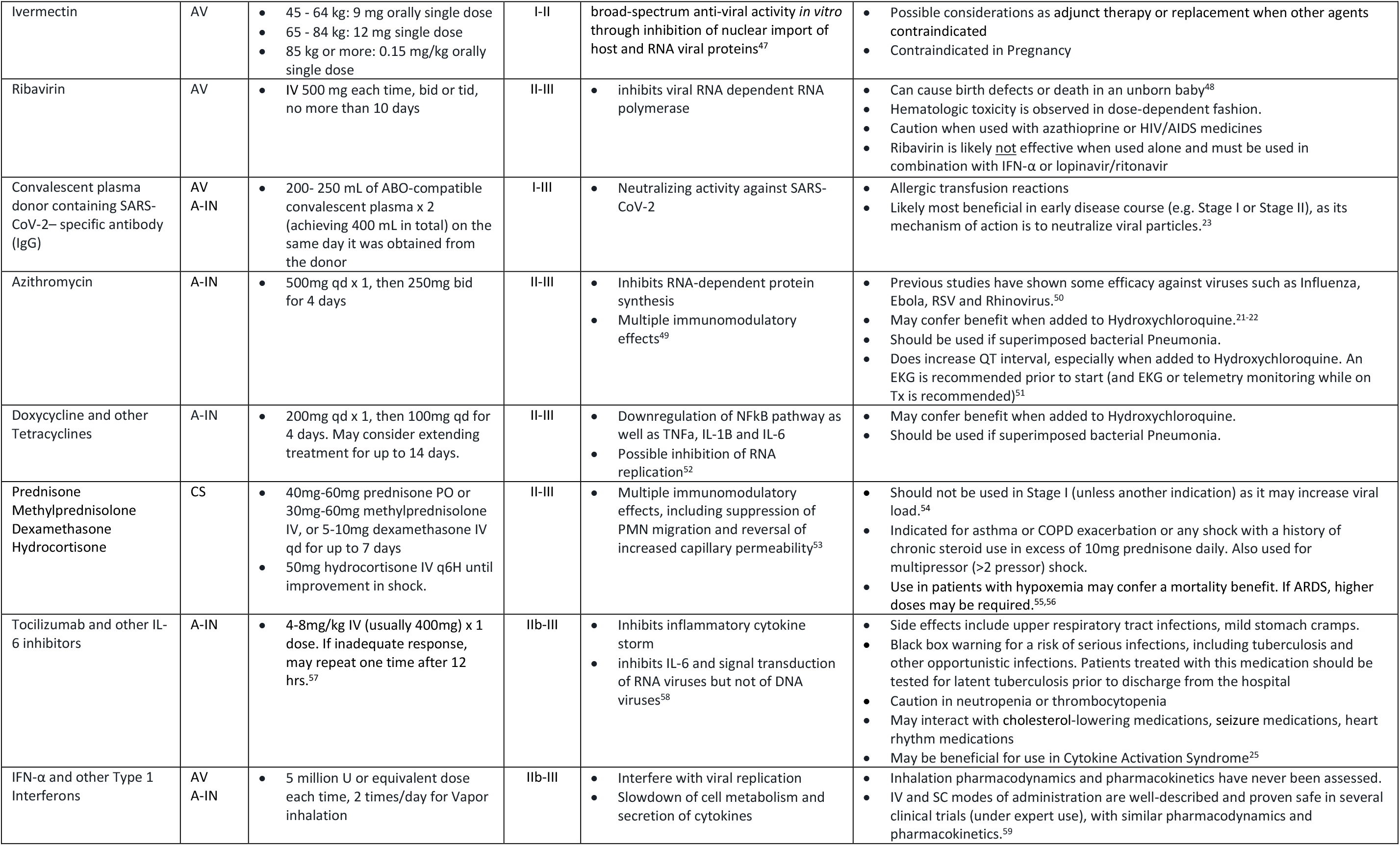
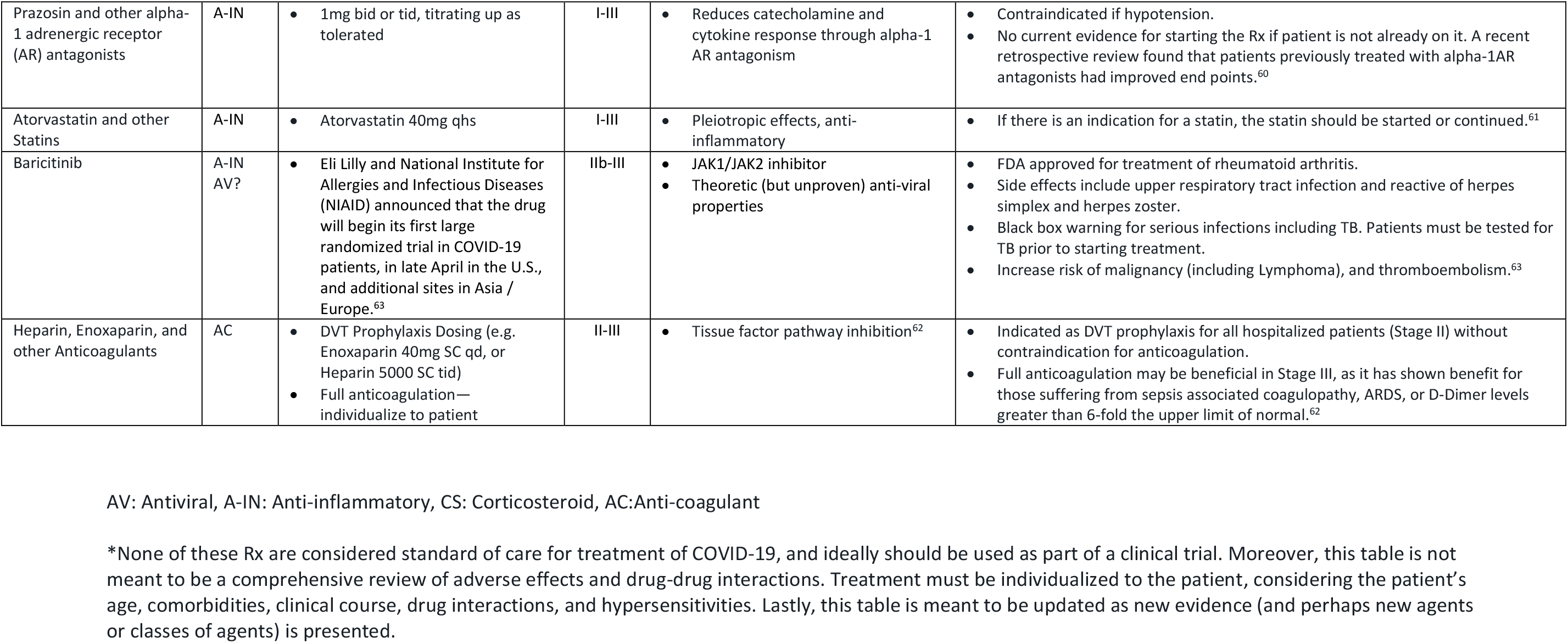
Summary of Investigational Treatments by COVD-19 Effect

| AGENT | EFFECT | DOSING | STAGE | MECHANISM | COMMENTARY |
| --- | --- | --- | --- | --- | --- |
| Remdesivir | AV | 200 mg IV x1, followed by 100mg qd for 5-10 days | I-III | RNA polymerase inhibitor<sup>39</sup> | Adverse effects include elevated ALT and AST, phlebitis, constipation, headache, nausea.<br>Theoretical risk of renal injury.<br>Should not be used in pregnancy due to lack of data.<br>Has limited drug-drug interactions (no significant CYP effect)<br>Clinical trials underway in the US, UK, and China<br>Showed efficacy in COVID-19 treatment<sup>40</sup> |
| Lopinavir/Ritonavir | AV | 200 mg/50 mg/capsule, 2 capsules PO bid for no more than 10-14 days | I-III | Protease inhibitor.<sup>41</sup> | Nearly 14% of patients cannot complete a course due to GI side effects.<sup>36</sup><br>Ritonavir is a potent CYP3A4 inhibitor (interacting with Rx such as apixaban, tacrolimus, and amiodarone).<br>In rare cases, Lopinavir/Ritonavir can cause liver injury, pancreatitis and cardiac toxicity.<br>Treatment in mostly Stage II-III patients was not found to be superior to standard of care. Subgroup analysis suggestive that earlier treatment (Stage I) might be beneficial.<sup>41</sup> |
| Favipiravir | AV | 1600mg PO bid x1d, then 600mg PO bid for up to 14 days | I-III | Broad spectrum inhibitor of RNA-dependent RNA polymerase<sup>42-43</sup> | Increases liver function parameters (AST, ALT and total bilirubin)<br>Testis toxicity and has a risk for teratogenicity and embryotoxicity<br>Was found to be superior to Lopinavir/Ritonavir in a small controlled study.<sup>44</sup> |
| Umifenovir | AV | 200mg q8h for up to 14 days | I-III | S protein / ACE-2 membrane fusion inhibitor<sup>45</sup> | Metabolism by CYP3A4. Caution with strong inhibitors or inducers.<br>Hypersensitivity risk increases in children under 2 years of age<br>Limited clinical evidence shows promise in COVID-19.<sup>46</sup> |
| Hydroxychloroquine | AV<br>A-IN | Stage I-II—400mg PO bid for first day followed by 200mg bid daily for 5 days<sup>20</sup><br>Stage III—May consider extending treatment (200mg bid) for up to 14 days.<sup>21,20</sup> | I-III | AV: replication-neutralization of the pH cellular organelles for gene replication<br>A-IN: inhibition of macrophage activation and reducing release of tissue TNF-α, IL-1, IL-6<sup>31-33</sup><br>A-IN: interfere with lysosomal activity and autophagy, disrupt membrane stability, alter signaling and transcriptional activity, which can then inhibit immune activation and cytokine production<sup>32</sup> | Adverse effects include: rash, nausea, and diarrhea. GI symptoms can be mitigated by taking with water; use with caution in diabetic patients may cause hypoglycemia.<sup>34</sup><br>Increased risk of retinopathy with a recommended maximal daily dose of 5.0 mg/kg. Avoid if history of retinal disease, macular degeneration, or previous treatment with tamoxifen.<sup>35</sup><br>Caution in patient at risk for QT prolongation. EKG at baseline and following initiation is generally advised, particularly in critically ill patients.<br>Contraindicated with Epilepsy, Porphyria, G6PD, and Myasthenia Gravis<br>Ideally used for patients hospitalized with Pneumonia<sup>36</sup><br>Consider for patients at high risk for disease progression.<br>Pre-Exposure/Post-Exposure prophylaxis is being studied |
| Chloroquine | AV<br>A-IN | Stage I-II – 500 mg bid for 5 days<br>Stage III—may consider extending treatment for up to 10 days | I-III | A-IN: decrease secretion and/or receptor expression of cytokines such as TNF-α<sup>33,37</sup><br>Interfere with lysosomal activity and autophagy, similar to Hydroxychloroquine<sup>32</sup> | Has greater adverse event profile than Hydroxychloroquine and possibly less efficacy.<br>Most common symptoms include abdominal cramps, nausea, anorexia.<br>Can increase QTc and result in hematologic effects (including hemolysis with G6PD deficiency). Can cause retinal toxicity and hypoglycemia.<br>As with Hydroxychloroquine, additional clinical trial data is necessary to determine efficacy and safety in COVID-19 patients.<sup>38</sup> |
| Ivermectin | AV | 45 - 64 kg: 9 mg orally single dose<br>65 - 84 kg: 12 mg single dose<br>85 kg or more: 0.15 mg/kg orally single dose | I-II | broad-spectrum anti-viral activity *in vitro* through inhibition of nuclear import of host and RNA viral proteins<sup>47</sup> | Possible considerations as adjunct therapy or replacement when other agents contraindicated<br>Contraindicated in Pregnancy |
| Ribavirin | AV | IV 500 mg each time, bid or tid, no more than 10 days | II-III | inhibits viral RNA dependent RNA polymerase | Can cause birth defects or death in an unborn baby<sup>48</sup><br>Hematologic toxicity is observed in dose-dependent fashion.<br>Caution when used with azathioprine or HIV/AIDS medicines<br>Ribavirin is likely <u>not</u> effective when used alone and must be used in combination with IFN-α or lopinavir/ritonavir |
| Convalescent plasma donor containing SARS-CoV-2– specific antibody (IgG) | AV<br>A-IN | 200- 250 mL of ABO-compatible convalescent plasma x 2 (achieving 400 mL in total) on the same day it was obtained from the donor | I-III | Neutralizing activity against SARS-CoV-2 | Allergic transfusion reactions<br>Likely most beneficial in early disease course (e.g. Stage I or Stage II), as its mechanism of action is to neutralize viral particles.<sup>23</sup> |
| Azithromycin | A-IN | 500mg qd x 1, then 250mg bid for 4 days | II-III | Inhibits RNA-dependent protein synthesis<br>Multiple immunomodulatory effects<sup>49</sup> | Previous studies have shown some efficacy against viruses such as Influenza, Ebola, RSV and Rhinovirus.<sup>50</sup><br>May confer benefit when added to Hydroxychloroquine.<sup>21-22</sup><br>Should be used if superimposed bacterial Pneumonia.<br>Does increase QT interval, especially when added to Hydroxychloroquine. An EKG is recommended prior to start (and EKG or telemetry monitoring while on Tx is recommended)<sup>51</sup> |
| Doxycycline and other Tetracyclines | A-IN | 200mg qd x 1, then 100mg qd for 4 days. May consider extending treatment for up to 14 days. | II-III | Downregulation of NFkB pathway as well as TNFa, IL-1B and IL-6<br>Possible inhibition of RNA replication<sup>52</sup> | May confer benefit when added to Hydroxychloroquine.<br>Should be used if superimposed bacterial Pneumonia. |
| Prednisone<br>Methylprednisolone<br>Dexamethasone<br>Hydrocortisone | CS | 40mg-60mg prednisone PO or 30mg-60mg methylprednisolone IV, or 5-10mg dexamethasone IV qd for up to 7 days<br>50mg hydrocortisone IV q6H until improvement in shock. | II-III | Multiple immunomodulatory effects, including suppression of PMN migration and reversal of increased capillary permeability<sup>53</sup> | Should not be used in Stage I (unless another indication) as it may increase viral load.<sup>54</sup><br>Indicated for asthma or COPD exacerbation or any shock with a history of chronic steroid use in excess of 10mg prednisone daily. Also used for multipressor (>2 pressor) shock.<br>Use in patients with hypoxemia may confer a mortality benefit. If ARDS, higher doses may be required.<sup>55,56</sup> |
| Tocilizumab and other IL-6 inhibitors | A-IN | 4-8mg/kg IV (usually 400mg) x 1 dose. If inadequate response, may repeat one time after 12 hrs.<sup>57</sup> | IIb-III | Inhibits inflammatory cytokine storm<br>inhibits IL-6 and signal transduction of RNA viruses but not of DNA viruses<sup>58</sup> | Side effects include upper respiratory tract infections, mild stomach cramps.<br>Black box warning for a risk of serious infections, including tuberculosis and other opportunistic infections. Patients treated with this medication should be tested for latent tuberculosis prior to discharge from the hospital<br>Caution in neutropenia or thrombocytopenia<br>May interact with cholesterol-lowering medications, seizure medications, heart rhythm medications<br>May be beneficial for use in Cytokine Activation Syndrome<sup>25</sup> |
| IFN-α and other Type 1 Interferons | AV<br>A-IN | 5 million U or equivalent dose each time, 2 times/day for Vapor inhalation | IIb-III | Interfere with viral replication<br>Slowdown of cell metabolism and secretion of cytokines | Inhalation pharmacodynamics and pharmacokinetics have never been assessed.<br>IV and SC modes of administration are well-described and proven safe in several clinical trials (under expert use), with similar pharmacodynamics and pharmacokinetics.<sup>59</sup> |
| Prazosin and other alpha-1 adrenergic receptor (AR) antagonists | A-IN | 1mg bid or tid, titrating up as tolerated | I-III | Reduces catecholamine and cytokine response through alpha-1 AR antagonism | Contraindicated if hypotension.<br>No current evidence for starting the Rx if patient is not already on it. A recent retrospective review found that patients previously treated with alpha-1AR antagonists had improved end points.<sup>60</sup> |
| Atorvastatin and other Statins | A-IN | Atorvastatin 40mg qhs | I-III | Pleiotropic effects, anti-inflammatory | If there is an indication for a statin, the statin should be started or continued.<sup>61</sup> |
| Baricitinib | A-IN<br>AV? | Eli Lilly and National Institute for Allergies and Infectious Diseases (NIAID) announced that the drug will begin its first large randomized trial in COVID-19 patients, in late April in the U.S., and additional sites in Asia / Europe.<sup>63</sup> | IIb-III | JAK1/JAK2 inhibitor<br>Theoretic (but unproven) anti-viral properties | FDA approved for treatment of rheumatoid arthritis.<br>Side effects include upper respiratory tract infection and reactive of herpes simplex and herpes zoster.<br>Black box warning for serious infections including TB. Patients must be tested for TB prior to starting treatment.<br>Increase risk of malignancy (including Lymphoma), and thromboembolism.<sup>63</sup> |
| Heparin, Enoxaparin, and other Anticoagulants | AC | DVT Prophylaxis Dosing (e.g. Enoxaparin 40mg SC qd, or Heparin 5000 SC tid)<br>Full anticoagulation—individualize to patient | II-III | Tissue factor pathway inhibition<sup>62</sup> | Indicated as DVT prophylaxis for all hospitalized patients (Stage II) without contraindication for anticoagulation.<br>Full anticoagulation may be beneficial in Stage III, as it has shown benefit for those suffering from sepsis associated coagulopathy, ARDS, or D-Dimer levels greater than 6-fold the upper limit of normal.<sup>62</sup> |

AV: Antiviral, A-IN: Anti-inflammatory, CS: Corticosteroid, AC:Anti-coagulant
\*None of these Rx are considered standard of care for treatment of COVID-19, and ideally should be used as part of a clinical trial. Moreover, this table is not meant to be a comprehensive review of adverse effects and drug-drug interactions. Treatment must be individualized to the patient, considering the patient's age, comorbidities, clinical course, drug interactions, and hypersensitivities. Lastly, this table is meant to be updated as new evidence (and perhaps new agents or classes of agents) is presented.

### Comorbidity

Data exists for early identification of cases at high risk of progression to severe COVID-19. One promising model created in China found that patients who developed severe COVID-19 possessed one of the following diseases: hypertension, diabetes, coronary heart disease, chronic respiratory disease, or tuberculosis. The same model cited age and various serological indicators (such as C-reactive protein (CRP), lactate dehydrogenase (LDH), bilirubin, and others) as factors associated with worse outcomes.^65^ Additional research confirmed, in a case-control study, that subjects with high Sequential Organ Failure Assessment (SOFA) scores, with age greater than 65, with hypertension, diabetes, and/or coronary heart disease were at greatest risk.^15^ Lastly, Research focusing on viral load and survival found that higher initial viral load is independently associated with worse prognosis.^2^

### Disease Progression

The most common presenting symptoms are fever and cough, followed by myalgia and fatigue. Less commonly, patients may present with sputum production, headache, or abdominal symptoms like diarrhea.^26^ In terms of disease progression, a case study of the first five patients diagnosed with COVID-19 in Europe points the way to two different clinical evolutions of the disease: 1. Presenting few symptoms, but showing high viral load from the respiratory tract; 2. A two-step disease process, with worsening of symptoms around 10 days of symptom onset in spite of decreased viral load in respiratory samples. In our model we plot the disease progression as a function of infection, survivability, and inflammation (Figure 2). We identify the inflection point where survival decreases as inflammation increases— approximately day 10 from symptom onset. Support for this is found in research by Chen J, Qi T, Liu L, et al. published in *The Journal of Infection*.^31^ Their research found that sepsis and ARDS in hospitalized patients starts around day 10 and 11, respectively. They also found temporal changes in inflammatory laboratory markers beginning at day 4 of illness onset. These included temporal changes in D-dimer, IL-6, serum ferritin, high-sensitivity cardiac troponin I, and lactate dehydrogenase. The differences were statistically significant between survivors and non-survivors for all time points. Figure 4 provides the percent change between survivors and non-survivors from day 4. In addition, Yang et al.^66^ found that the patients admitted to the ICU with severe hypoxemia had a 50% probability of survival at day 7 of ICU admission (corresponding to Day ∼17 in Figure 2).

**Figure 4.**
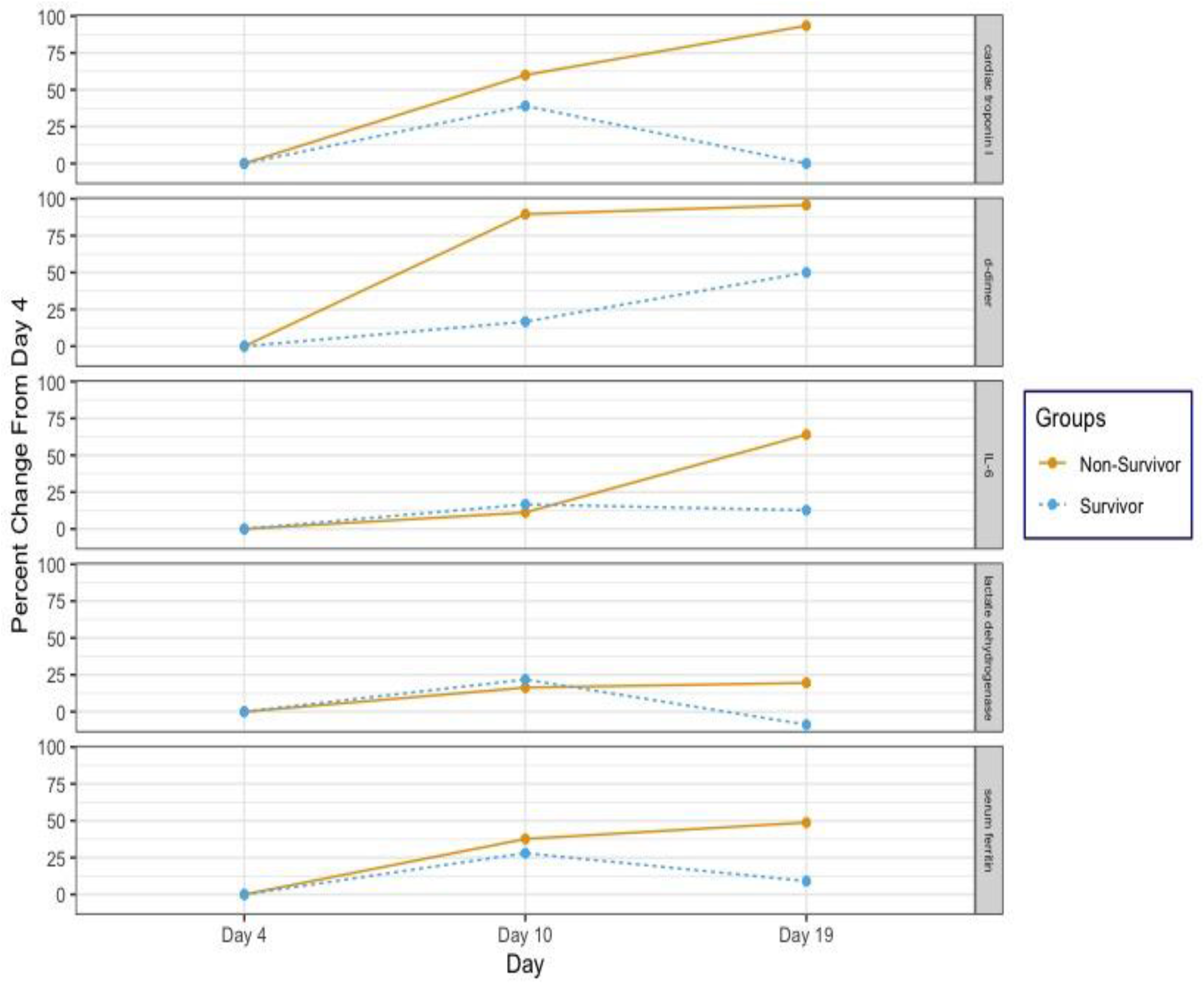
Percent Change in Clinical Measures between Survivors and Non-Survivors Source: Chen J, Qi T, Liu L, et al. Clinical progression of patients with COVID-19 in Shanghai, China. [published online March 19, 2020]. *J Infect*. 2020;S0163-4453(20)30119-5. doi:10.1016/j.jinf.2020.03.004.

### Stage I

The incubation period is on average 5 days. In most patients, initial presenting symptoms are mild (though a small number of patients can be asymptomatic throughout the course of the disease). Stage I symptoms include fever, cough, fatigue, and body aches, but, in a minority of cases can include headache, GI symptoms, anosmia, as well as others. Duration of initial symptoms is 5-7 days, correlating with a peak in viral load.^26^ During this time, the appropriate diagnostic test is a nasopharyngeal PCR. Laboratory studies may include an elevated D-Dimer and prothrombin time, as well as lymphopenia (see Figure 2). Given that symptoms in this stage are mild, and correlated with viremia, the appropriate treatment modality is supportive care or an antiviral. Nevertheless, treatment must be individualized, based on a patient’s age, comorbidities, presenting symptoms, and drug interactions (see Figure 3 and Table 1).

### Stage II

Some patients progress into Stage II, which is characterized by a decrease in viral levels and an increase in inflammation that initially localizes to the lungs. Infiltrates are typically seen on chest x-ray (CXR) or computed tomography (CT). Similar to symptom duration in Stage I, the typical symptom course in Stage II is also 5-7 days. Treatment with antivirals is still indicated, but given an average decrease in viral levels during this stage, that treatment is theoretically less effective than in Stage I. Moreover, Stage II is divided into two sub-stages (IIA and IIB), depending on whether a patient is hypoxemic or not. This distinction is important for management (see Figure 2). In Stage IIB, the patient is significantly dyspneic and may benefit, depending on age and comorbidities, from the use of corticosteroids or other anti-inflammatory treatments (see Figure 3).

### Stage III

Although only a minority of patients (estimated at 10-15%) progress to Stage III, mortality within this stage is considerable (estimated at 20-30%). The morbidity and mortality are generally due to uncontrolled inflammation, which at this point is systemic. The most important symptom is respiratory distress (correlating in a typical patient to a Pulse Ox ≤ 92%). Laboratory markers include significantly increased CRP and IL-6 levels.^66-67^ As in Stage II, treatment may include anti-virals (if the patient is still viremic), but agents to counteract inflammation and its effects (such as microthrombi) must be considered (see Figure 2). A summary of investigational therapies can be found in Table 1.

### Pre-Exposure and Post-Exposure Prophylaxis

A number of clinical trials are exploring pre-exposure and post-exposure prophylaxis. There is no definitive evidence that any particular treatment modality is effective but antivirals, hydroxychloroquine, and convalescent plasma are being proposed. Anti-virals, like Remdesivir, may proof beneficial at any stage of disease.^39-40^ Hydroxychloroquine is postulated to have anti-viral properties, and it has been definitively proven as an immunomodulator.^32^ Convalescent plasma provides the antibody support needed to envelope and destroy the virus while preventing the exuberant immune response or cytokine release that leads to significant pathology, particularly in Stages IIb and III.^23^

## Limitations

This review has several limitations. First, the incredible volume and speed at which data is published about the treatment of COVID-19 indicates that research findings and recommendations may change. Second, the research used to create the review came from small studies, often-times with very few controls. Third, the articles were limited to English-language publications or translations, so relevant international data could be lacking.

## Conclusion

This paper presents the first evidence-based recommendations for individualized treatment for COVID-19. Based upon the observed transmission and mortality rates, health professionals urgently need to align patient baseline risk to disease stage and investigational treatment options. The COVID-19 pandemic represents the greatest public health crisis in three generations; the need for comprehensive management cannot be overstated.

## Data Availability

Our study was based on a review of existing sources.

## References

1. Heyn PC, Meeks S, Pruchno R. Methodological guidance for a quality review article. Gerontologist. 2019 Mar 14;59(2):197–201. https://doi.org/10.1093/geront/gny123

2. Centers for Disease Control and Prevention. Cases in U.S. 2020, April 14. https://www.cdc.gov/coronavirus/2019-ncov/cases-updates/cases-in-us.html#2019coronavirus-summary. Accessed April 14, 2020

3. The Institute for Health Metrics and Evaluation. COVID-19 projections assuming full social distancing through May 2020. https://covid19.healthdata.org/united-states-of-america. Accessed April 9, 2020.

4. Worldometers.info. COVID-19 coronavirus pandemic. https://www.worldometers.info/coronavirus/. Accessed April 5, 2020.

5. Craven M, Liu L, Mysore M, et al. COVID-19: briefing note, March 9, 2020. McKinsey and Company. https://www.mckinsey.com/~/media/McKinsey/Business%20Functions/Risk/Our%20Insights/COVID%2019%20Implications%20for%20business/COVID%2019%20March%209/COVID-19-Briefing-note-March-9-2020-v5.ashx.. Accessed April 6, 2020.

6. Biggerstaff M, Cauchemez S, Reed C, et al. Estimates of the reproduction number for seasonal, pandemic, and zoonotic influenza: a systematic review of the literature. BMC Infect Dis. 2014;480. doi:https://doi.org/10.1186/1471-2334-14-480.

7. Su S, Wong G, Shi W, et al. Epidemiology, genetic recombination, and pathogenesis of coronaviruses. Trends in Microbiology. 2016;24(6):490–502. doi:http://dx.doi.org/10.1016/j.tim.2016.03.003

8. Chen N, Zhou M, Dong X, et al. Epidemiological and clinical characteristics of 99 cases of 2019 novel coronavirus pneumonia in Wuhan, China: a descriptive study. Lancet. 2020;395(10223):507–513. doi:https://doi.org/10.1016/S0140-6736(20)30211-7

9. Arentz M, Yim E, Klaff L, et al. Characteristics and outcomes of 21 critically ill patients with COVID-19 in Washington state. JAMA. [published online March 19, 2020]. doi:10.1001/jama.2020.4326

10. Guan WJ, Ni ZY, Liang WH, et al. Clinical characteristics of coronavirus disease 2019 in China. N Engl J Med. [published online February 28, 2020]. doi:10.1056/NEJMoa2002032

11. Siddiqi HS, Mehra MR. COVID-19 illness in native and immunosuppressed states: a clinical therapeutic staging proposal. J Heart Lung Transplant. [published online March 20, 2020]. doi:10.1016/j.healun.2020.03.012

12. Parker BS, Walker KH. Clinical course, prognosis, and epidemiology. Brigham and Women’s Hospital COVID-19 Clinical Guidelines. Retrieved from https://covidprotocols.org/protocols/01-clinical-course-prognosis-and-epidemiology. Accessed April 5, 2020.

13. Marik P. EVMS critical care COVID-19 management protocol. Eastern Virginia Medical School, Norfolk, VA. April 6, 2020. https://www.evms.edu/media/evms_public/departments/internal_medicine/EVMS_Critical_Care_COVID-19_Protocol.pdf

14. Hsieh YH, Lee JY, Chang HL. SARS Epidemiology modeling. Emerg Infect Dis. 2004;10(6):1165–1167. doi:10.3201/eid1006.031023

15. Zhou F, Yu T, Du R, et al. Clinical course and risk factors for mortality of adult inpatients with COVID-19 in Wuhan, China: a retrospective cohort study. Lancet. 2020;395(10229):1054–1062. doi:10.1016/s0140-6736(20)30566-3

16. Yang X, Yu Y, Xu J, et al. Clinical course and outcomes of critically ill patients with SARS- Co-V-2 pneumonia in Wuhan, China: a single-centered, retrospective, observational study. Lancet Respir. Med. [published online February 24, 2020]. doi:https://doi.org/10.1016/S2213-2600(20)30079-5

17. Hsueh PR, Huang LM, Chen PJ, Kao CL, Yang PC. Chronological evolution of IgM, IgA, IgG and neutralisation antibodies after infection with SARS-associated coronavirus. Clin Microbiol Infect. 2004;10(12):1062–1066. doi:10.1111/j.1469-0691.2004.01009.x

18. Ware LB, Matthay MA. The acute respiratory distress syndrome. N Engl J Med. 2000;342(18):1334–1349.

19. Savarino A, Boelaert JR, Cassone A, Majori G, Cauda R. Effects of chloroquine on viral infections: an old drug against today’s diseases? Lancet Infect Dis. 2003;3(11):722–727.

20. Yao X, Ye F, Zhang M, et al. In vitro antiviral activity and projection of optimized dosing design of hydroxychloroquine for the treatment of severe acute respiratory syndrome coronavirus 2 (SARS-CoV-2). Clin Infect Dis. [published online March 9, 2020]. doi:10.1093/cid/ciaa237

21. Gautret P, Lagier JC, Parola P, et al. Hydroxychloroquine and azithromycin as a treatment of COVID-19: results of an open-label non-randomized trial. Int J Antimicrob Agents. [published online March 20, 2020]. doi:10.1016/j.ijantimicag.2020.105949

22. Gautret P, Lagier JC, Parola P, et al. Clinical and microbiological effect of a combination of hydroxychloroquine and azithromycin in 80 COVID-19 patients with at least a six-day follow up: a pilot observational study. Travel Medicine and Infectious Disease. [published online April 11, 2020]. doi:10.1016/j.tmaid.2020.101663

23. Shen C, Wang Z, Zhao F, et al. Treatment of 5 critically ill patients with COVID-19 with convalescent plasma. JAMA. [published online March 27, 2020]. doi:10.1001/jama.2020.4783

24. Chen RC, Tang XP, Tan SY, et al. Treatment of severe acute respiratory syndrome with glucosteroids: the Guangzhou experience. Chest. 2006, 129(6):1441–1452. doi:10.1378/chest.129.6.1441

25. Xu X, Han M, Li T, et al. Effective treatment of severe COVID-19 patients with tocilizumab. [published online March 5, 2020]. ChinaXiv. doi:10.12074/202003.00026

26. Huang C, Wang Y, Li X, et al. Clinical features of patients infected with 2019 novel coronavirus in Wuhan, China. Lancet. 2020;395(10223):497–506. doi:10.1016/S0140-6736(20)30183-5.

27. Wang D, Hu B, Hu C, et al. Clinical characteristics of 138 hospitalized patients with 2019 novel coronavirus-infected pneumonia in Wuhan, China. JAMA. [published online February 8, 2020.] doi:10.1001/jama.2020.1585

28. Force ADT, Ranieri VM, Rubenfeld GD, et al. Acute respiratory distress syndrome: The Berlin Definition. JAMA. 2012;307(23):2526–2533. doi:10.1001/jama.2012.5669

29. Riviello Ed, Kiviri W, Twagirumugabe T, et al. Hospital incidence and outcomes of the acute respiratory distress syndrome using the Kigali modification of the Berlin Definition. Am J. Respir Crit Care Med. 2016;193(1):52–59. doi:10.1164/frccm.201503-0584OC

30. People Who Are at Higher Risk for Severe Illness. (2020, April). Retrieved April 13, 2020, from https://www.cdc.gov/coronavirus/2019-ncov/need-extra-precautions/groups-at-higher-risk.html

31. Chen J, Qi T, Liu L, et al. Clinical progression of patients with COVID-19 in Shanghai, China. [published online March 19, 2020]. J Infect. 2020;S0163-4453(20):30119-30125. doi:10.1016/j.jinf.2020.03.004

32. Schrezenmeier E, Dörner T. Mechanisms of action of hydroxychloroquine and chloroquine: implications for rheumatology. Nat Rev Rheum. 2020;16(3):155–166.

33. Sahraei Z, Shabani M, Shokouhi S, Saffaei A. Aminoquinolines against coronavirus disease 2019 (COVID-19): chloroquine or hydroxychloroquine. Int J Antimicrob Agents. [published online March 17, 2020] doi:10.1016/j.ijantimicag.2020.105945

34. Bethel M, Yang FM, Li S, et al. Hydroxychloroquine in patients with systemic lupus erythematosus with end-stage renal disease. J Investig Med. 2016;64(4):908–910.

35. April J, Ung C, Young L, et al. Hydroxychloroquine retinopathy—implications of research advances for rheumatology care. Nat Rev Rheum. 2018;14(12):693–703.

36. Bhimraj A, Morgan RL, Shumaker AH, et al. Infectious Diseases Society of America guidelines on the treatment and management of patients with COVID-19. [published online April 11, 2020]. https://www.idsociety.org/globalassets/idsa/practice-guidelines/covid-19/treatment/idsa-covid-19-gl-tx-and-mgmt-4-11-20-1058-am-edt.pdf

37. van den Borne BE, Dijkmans BA, de Rooij HH, le Cessie S, Verweij CL. Chloroquine and hydroxychloroquine equally affect tumor necrosis factor-alpha, interleukin 6, and interferon-gamma production by peripheral blood monomuclear cells. J Rheumatol. 1997;24(1):55–60. https://europepmc.org/article/med/9002011

38. Pharmacists Advancing Healthcare. Assessment of evidence for COVID-19-related treatments. Retrieved from https://www.ashp.org/-/media/assets/pharmacy-practice/resource-centers/Coronavirus/docs/ASHP-COVID-19-Evidence-Table.ashx

39. Wang M, Cao R, Zhang L, et al. Remdesivir and chloroquine effectively inhibit the recently emerged novel coronavirus (2019-nCoV) in vitro. Cell Res. 2020;30;269–271. doi:https://doi.org/10.1038/s41422-020-0282-0

40. Grein J, Ohmagari N, Shin D, et al. Compassionate use of remdesivir for patients with severe Covid-19. [published online April 10, 2020]. NEJM. doi:10.1056/NEJMoa2007016

41. Cao B, Wang Y, Wen D, et al. A trial of lopinavir-ritonavir in adults hospitalized with severe Covid-19. [published online March 18, 2020]. NEJM. doi:10.1056/NEJMoa2001282

42. Furuta Y, Komeno T, Nakamura T. Favipiravir (T-705), a broad spectrum inhibitor of viral RNA polymerase. Proc Jpn Acad Ser B Phys Biol Sci. 2017;93(7), 449–463. doi:https://doi.org/10.2183/pjab.93.027

43. Nagata T, Lefor AK, Hasegawa M, Ishii M. Favipiravir: a new medication for the Ebola virus disease pandemic. Disaster Medicine and Public Health Preparedness. 2015;9(1):79–81. doi:10.1017/dmp.2014.151

44. Qingxian C, Yang M, Liu D, et al. Experimental treatment with Favipiravir for COVID-19: an open-label control study. [published online March 1, 2020]. Engineering. doi:10.1016/j.eng.2020.03.007

45. Kadam RU, Wilson IA. Structural basis of influenza virus fusion inhibition by the antiviral drug Arbidol. Proc Natl Acad Sci U S A. 2017:114(2):206–214. doi:10.1073/pnas.1617020114

46. Wang Z, Yang B, Li Q, Wen L, Zhang R. Clinical features of 69 cases with coronavirus disease 2019 in Wuhan, China. Clin Infect Dis. [published online March 16, 2020]. doi:10.1093/cid/ciaa272

47. Cally L, Druce JD, Catton MG. The FDA-approved drug Ivermectin inhibits the replication of SARS-CoV-2 in vitro. [published online April 3, 2020]. Antiviral Res. doi:10.1016/j.antiviral.2020.104787

48. Roberts SS, Miller RK, Jones JK, et al. The Ribavirin pregnancy registry: findings after 5 years of enrollment, 2003-2009. Birth Defects Res A Clin Mol Teratol. 2010;88(7):551–559. doi:10.1002/bdra.20682

49. Ohe M, Shida H, Jodo S, et al. Macrolide treatment for COVID-19:will this be the way forward? Biosci Trends. [published online April 5, 2020] doi:10.5582/bst.2020.03058

50. Amsden GW. Anti-inflammatory effects of macrolides—an underappreciated benefit in the treatment of community-acquired respiratory tract infections and chronic inflammatory pulmonary conditions? Journal of Antimicrobial Chemotherapy. 2005;55(1):10–21. doi:https://doi.org/10.1093/jac/dkh519

51. Simpson TF, Kovacs RJ, Stecker EC. Ventricular arrhythmia risk due to hydroxychloroquine-azithromycin treatment for COVID-19. Cardiology Magazine. 2020, March 29. Retrieved from https://www.acc.org/latest-in-cardiology/articles/2020/03/27/14/00/ventricular-arrhythmia-risk-due-to-hydroxychloroquine-azithromycin-treatment-for-covid-19

52. Sodhi M, Etminan M. Therapeutic potential for tetracyclines in the treatment of COVID- 19. Pharmacotherapy. [published online April 8, 2020]. doi:10.1002/pharm2395

53. Puckett Y, Gabbar A, Bokhari AA. Prednisone. StatPearls. Retrieved from https://www.ncbi.nlm.nih.gov/books/NBK534809/ on April 2, 2020.

54. Lee N, Chan KCA, Hui DS, et al. Effects of early corticosteroid treatment on plasma SARS-associated Coronavirus RNA concentrations in adult patients. J Clin Virol. 2004;31(4):304–309. doi:10.1016/j.jvc.2004.07.006

55. Tang N, Bai H, Chen X, Gong J, Li D, Sun Z. Anticoagulant treatment is associated with decreased mortality in severe coronavirus disease 2019 patients with coagulopathy. Journal of Thrombosis and Haemostasis. [published online March 27, 2020]. https://doi.org/10.1111/jth.14817

56. Wu C, Chen X, Cai Y, et al. Risk factors associated with acute respiratory distress syndrome and death in patients with Coronavirus Disease 2019 pneumonia in Wuhan, China. JAMA Intern Med. [published online March 13, 2020]. doi:10.1001/jamainternmed.2020.0994

57. Walker KH, Pearson JC. Therapeutics and clinical trials. Brigham and Women’s Hospital: COVID-19 Clinical Guidelines. Retrieved from https://covidprotocols.org/protocols/04-therapeutics-and-clinical-trials on April 11, 2020.

58. Ruan Q, Yang K, Wang W, Jiang L, Song J. Clinical predictors of mortality due to COVID- 19 based on an analysis of data of 150 patients from Wuhan, China. Intensive Care Med. [published online March 3, 2020]. doi:10.1007/s00134-020-05991-x

59. Sallard E, Lescure FX, Yazdanpanah Y, Mentre F, Peiffer-Smadja N. Type 1 interferons as a potential treatment against COVID-19. Antiviral Res. [published online April 7, 2020]. doi:10.1016/j.antiviral.2020.104791

60. Konig MF, Powell M, Staedtke V, et al. Targeting the catecholamine-cytokine axis to prevent SARS-CoV-2 cytokine storm syndrome. MedRxiv. [published online April 8, 2020]. Doi:https://doi.org/10.1101/2020.04.02.20051565

61. Massachusetts General Hospital. Massachusetts General Hospital COVID-19 treatment guidance. Retrieved from https://www.massgeneral.org/assets/MGH/pdf/news/coronavirus/mass-general-COVID-19-treatment-guidance.pdf on April 13, 2020.

62. Tang N, Bai H, Chen X, Gong J, Li D, Sun Z. Anticoagulant treatment is associated with decreased mortality in severe coronavirus disease 2019 patients with coagulopathy. Journal of Thrombosis and Haemostasis. [published online March 27, 2020]. https://doi.org/10.1111/jth.14817

63. Eli Lilly. Lilly begins clinical testing of therapies for COVID-19. Retrieved from https://investor.lilly.com/news-releases/news-release-details/lilly-begins-clinical-testing-therapies-covid-19 on April 13, 2020.

64. Gong J, Ou J, Qiu X, et al. A tool to early predict severe 2019-novel coronavirus pneumonia (COVID-19): a multicenter study using the risk nomogram in Wuhan and Guangdong, China. MedRxiv. [published online March 20, 2020]. doi:https://doi.org/10.1101/2020.03.17.20037515

65. Lescure FX, Bouadma L, Nguyen D, et al. Clinical and virological data of the first cases of COVID-19 in Europe: a case series. Lancet Infect Dis. [published online March 22, 2020]. doi:https://doi.org/10.1016/S1473-3099(20)30200-0

66. Yang X, Yu Y, Xu J, et al. Clinical course and outcomes of critically ill patients with SARS- Co-V-2 pneumonia in Wuhan, China: a single-centered, retrospective, observational study. Lancet. [published online February 24, 2020]. doi:https://doi.org/10.1016/S2213-2600(20)30079-5

67. Zhang C, Wu Z, Li JW, Zhao H, Wang GQ. The cytokine release syndrome (CRS) of severe COVID-19 and Interleukin-6 receptor (IL-6R) antagonist Tocilizumab may be the key to reduce the mortality [published online ahead of print, 2020 Mar 29]. Int J Antimicrob Agents. 2020;105954. doi:10.1016/j.ijantimicag.2020.105954

